# Knowledge–Attitude Gap and Unsupervised Use of Artificial Intelligence Tools Among Swiss Primary Care Physicians: A Multicentric Cross-Sectional Survey

**DOI:** 10.1101/2025.03.22.25324458

**Authors:** Marco Vecellio

**Affiliations:** Specialist in General Internal Medicine and Psychosomatic Medicine, mediX Group Practice, Hegibachstrasse 2, 8032 Zurich, Switzerland

**Keywords:** artificial intelligence, primary care, general practitioners, physician readiness, knowledge–attitude gap, implementation science, patient safety, Switzerland, survey research, managed care

## Abstract

**Background:** Adoption of artificial intelligence (AI) by physicians is accelerating; among United States physicians self-reported use rose from 38% to 66% between 2023 and 2024. Implementation strategies in primary care have typically framed physician resistance as the principal barrier, but emerging evidence points to a different and arguably more pressing pattern: enthusiastic clinical use of AI by physicians who themselves report inadequate knowledge to use it safely. The magnitude of this pattern in organised primary care has not been quantified.

**Methods:** We conducted a closed-network, census-style online survey (August–September 2024) within mediX Switzerland, addressing all 620 primary care physicians on the combined electronic distribution lists of the four consenting subnetworks (mediX Zurich, Bern, Romandie, Ticino; approximately 70% of the full network of >900 physicians). 160 physicians responded (25.8%) and 155 met the inclusion criterion of completing at least 2 of 4 core readiness items. Analyses used Wilson Score 95% confidence intervals (CI), Cohen’s *h* and Cohen’s *g* effect sizes (Cohen 1988), an exact McNemar test for the paired primary-outcome comparison, Mann–Whitney *U* with rank-biserial *r* for subgroup contrasts, a knowledge-threshold sensitivity analysis (level ≥ 4 vs ≥ 3) and Manski-style non-response bounds against the sampling frame. Reporting follows STROBE and CHERRIES.

**Results:** 107/155 (69.0%, 95% CI 61.4–75.8) reported positive attitudes toward AI; 23/155 (14.8%, 95% CI 10.1–21.3) reported adequate self-rated knowledge — a 54.2-percentage-point between-marginal gap (Cohen’s *h* = 1.17), confirmed by the paired McNemar test (87 vs 3 discordant pairs, *p* < 0.001; Cohen’s *g* = 0.542). 43/155 (27.7%, 95% CI 21.3–35.3) already used large-language-model tools clinically; 127/155 (81.9%, 95% CI 75.1–87.2) endorsed training. Endorsement of nine prespecified implementation domains formed three descriptive support tiers (≥ 70%: administrative support, image analysis; 50–69%: medication, diagnostic, therapy support; < 50%: prevention, complex case, research, patient communication).

**Conclusions:** Among AI-engaged Swiss primary care physicians, the dominant practical risk is unsupervised adoption rather than resistance. Educational and governance interventions appear required *before* further deployment.

## 1. Introduction

Artificial intelligence (AI) is moving from experimental setting into everyday clinical use at a pace that few healthcare systems have organised themselves to absorb. Among United States physicians, self-reported AI use rose from 38% in 2023 to 66% in 2024 [1]; technical capabilities have advanced from radiology-specific deep-learning models [2] to generalist medical foundation models [3] and large language models that exceed average human performance on standard medical examinations [4]. Primary care, situated at the entry point of most patient pathways, will absorb a substantial share of these tools — yet the discipline lacks structured baseline data on how prepared its physicians are to use them safely.

The dominant framing of AI implementation has emphasised physician *resistance* as the principal obstacle to overcome [5,6]. This framing organises both research and policy: knowledge gaps are treated as an information problem to be solved with continuing-medical-education campaigns; trust deficits are treated as a relationship problem to be solved with explainability features; regulatory caution is treated as a coordination problem to be solved with frameworks such as the European Union AI Act [7] and the World Health Organization guidance on large multi-modal models [8]. Each of these intervention classes presupposes that the binding constraint on safe adoption is *not yet* having physicians engaged with AI.

A different pattern, less often quantified, may now be more pressing. Recent UK survey data show that general practitioners are already using generative AI in clinical work in non-trivial proportions and frequently without institutional sanction or training [9]; a systematic review across primary care contexts identifies persistent knowledge–attitude gaps in which positive attitudes outpace measurable competence [6]. If this pattern holds in organised primary care networks elsewhere, then the binding constraint is no longer convincing reluctant physicians to engage with AI — it is *educating and governing physicians who are already using AI* and who themselves report inadequate competence to do so.

The Swiss primary care setting offers an instructive vantage point on this question. The mediX Switzerland network — a hybrid managed-care organisation comprising more than 900 primary care physicians across the German-, French- and Italian-speaking regions — combines coordinated care delivery with substantial physician autonomy, an organisational profile broadly comparable to Accountable Care Organisations in the United States [10] and to Primary Care Networks in the United Kingdom. To our knowledge, no quantitative readiness assessment within such a multilingual organised primary care network has yet been published.

This study therefore investigates physician readiness for clinical AI within mediX Switzerland with three specific aims: (i) to quantify the gap between self-rated knowledge and attitudes toward AI; (ii) to characterise the prevalence and pattern of current AI use in clinical practice relative to self-rated competence; and (iii) to elicit implementation priorities, perceived benefits and perceived barriers in a form usable for the design of educational and governance interventions.

## 2. Materials and Methods

### 2.1. Study design and setting

This was a cross-sectional, anonymous, web-based survey of primary care physicians within mediX Switzerland, conducted between 1 August and 30 September 2024. mediX is a non-profit cooperative network of independent primary care practices operating under capitated managed-care contracts with major Swiss health insurers; at the time of fieldwork it comprised approximately 900 physicians distributed across multiple regional subnetworks. Four of these subnetworks — mediX Zurich, mediX Bern, mediX Romandie and mediX Ticino, jointly representing the largest regional units across the German-, French- and Italian-speaking regions of Switzerland — formally consented to participation in this study and provided the corresponding electronic distribution lists. The remaining smaller subnetworks were not approached, primarily because of the absence of central distribution infrastructure or the limited time available within their administrative cycle. The four participating subnetworks together cover the three principal Swiss linguistic regions and combine elements of independent practice and coordinated care.

### 2.2. Participants, sampling frame and design classification

The sampling frame consisted of all 620 primary care physicians on the combined electronic distribution lists of the four consenting subnetworks (approximately 70% of the overall mediX network of >900 physicians). All 620 physicians received the same survey invitation by secure professional email (HIN, the dominant Swiss medical communications network), with two reminder emails at two-week intervals. No financial or material incentive was offered.

This sampling design is best characterised as a **closed-network, census-style invitation with self-selected respondents**: the invitation was directed at the *entire* eligible frame (a census of the four consenting subnetworks), but participation was voluntary, optional and uncompensated. Given the well-documented high workload of practising primary care physicians and the absence of incentives, a non-trivial element of self-selection toward AI-engaged physicians is to be expected; the 25.8% response rate is consistent with this expectation and with international physician-survey benchmarks (typically 20–35% for non-incentivised online instruments). For consistency with reporting-standard terminology, we therefore avoid the term «convenience sample», which would imply ad-hoc accessibility-based recruitment, in favour of the more precise designation above. Eligibility criteria were: practising primary care physician within the mediX network; (implicit) informed consent through completion of the questionnaire. There were no exclusion criteria beyond non-completion of the readiness items.

### 2.3. Survey instrument

The 35-item instrument (German master version; certified translations into French and Italian) was developed by the author from existing literature on physician AI attitudes [5,6,11,12], adapted to the Swiss context, and pilot-tested with five primary care physicians who were not subsequently invited to the main study. The pilot identified ambiguous wording in three items (subsequently revised) and confirmed an estimated completion time of 10–15 minutes. Domains covered: demographics and practice context (region, age band, gender, years of experience, practice size, urbanicity); self-rated AI knowledge (5-point scale, 1 = very low to 5 = excellent); attitudes toward clinical AI (5-point Likert, 1 = very negative to 5 = very positive); current personal use of large-language-model tools for medically-relevant work (yes/no with free-text field); perceived benefits (multiple choice, ≤ 5 of 9 prespecified items); perceived barriers (multiple choice, ≤ 5 of 10); implementation priorities (multiple choice, ≤ 5 of 9 clinical domains); framework conditions considered necessary; demand for training (5-point Likert). The full instrument in all three deployed languages (German, French, Italian) is openly available at the Zenodo repository accompanying this manuscript (DOI 10.5281/zenodo.20081687, CC BY 4.0); a bilingual (German/English) data codebook is provided as Supplementary File 1.

### 2.4. Outcomes

The pre-specified primary outcome was the **knowledge–attitude gap**, defined as the absolute difference between the proportion of respondents reporting positive attitudes toward AI (Likert categories 4–5) and the proportion reporting adequate self-rated knowledge (categories 4–5). Secondary outcomes were: prevalence of current clinical AI use, demand for training, ranked implementation priorities, perceived benefits and barriers, and required framework conditions. We pre-specified that “adequate knowledge” would be defined conservatively as level 4 or 5; a sensitivity analysis re-defining adequate knowledge as level 3 or higher was performed (see Section 2.6).

### 2.5. Statistical analysis

Analyses were performed in Python 3.11 with pandas 2.1 (BSD-3-Clause), scipy 1.11 (BSD-3-Clause), numpy 1.26 (BSD-3-Clause), openpyxl 3.1 (MIT) and matplotlib 3.8 (PSF; figure-generation only). The full analysis script (`analysis_swissAIsurvey_v11.py`, internally versioned to v12 to reflect the addition of paired-proportion analyses) is openly available at Zenodo, DOI 10.5281/ zenodo.20081687, under a Creative Commons Attribution 4.0 International (CC BY 4.0) licence; see Section 2.7 and the *Data Availability Statement*. 95% Wilson Score confidence intervals (CIs) [13] were computed for *all reported proportions and absolute proportion differences*, in preference to normal-approximation intervals because Wilson Score has superior coverage for moderate samples and proportions away from 0.5.

Effect sizes for *between-marginal* comparisons of two proportions were quantified using Cohen’s *h*, with the original Cohen 1988 cut-offs of 0.20 (small), 0.50 (medium) and 0.80 (large) [14]. We note explicitly that Cohen’s *h* was originally developed for *independent* proportions; in the present study the principal contrast (positive attitude vs. adequate self-rated knowledge) involves two proportions measured on the same 155 respondents and is therefore strictly a *paired* comparison. To address this methodological subtlety, the principal between-marginal *h* values are complemented by an exact McNemar test of the discordant 2 × 2 cells and by Cohen’s *g* for paired proportions (Cohen 1988 cut-offs 0.05 / 0.15 / 0.25) [14]. Group comparisons of ordinal items used the Mann–Whitney *U* test (smaller-rank-sum convention) with rank-biserial correlation as effect size. Spearman’s ρ is reported for ordinal–ordinal correlations (e.g., years of practice × attitude). No imputation of missing data was performed; per-item denominators are reported alongside each estimate.

### 2.6. Sensitivity analyses

Three sensitivity analyses were specified prior to data inspection. First, a **non-response bias analysis** computed Manski-style worst-case bounds for the population-level training-demand estimate against the full sampling frame (n = 620), assuming that among the 465 non-responders training demand was respectively (a) 0% (lower bound), (b) the same as among responders (responder-equivalent), or (c) 100% (upper bound). Second, a **knowledge-threshold sensitivity analysis** re-computed the primary outcome using level ≥ 3 (“moderate or higher”) as the threshold for adequate knowledge. Third, a **subgroup analysis** compared current AI users with non-users on attitude scores using the Mann–Whitney *U* test, reporting rank-biserial correlation as effect size.

### 2.7. Reporting standards

Reporting follows the Checklist for Reporting Results of Internet E-Surveys [15] and the Strengthening the Reporting of Observational Studies in Epidemiology (STROBE) statement for cross-sectional studies [16]; both completed checklists are appended (Supplementary Files S1 and S2). The participant flow diagram is provided as Supplementary Figure S3 in this manuscript.

### 2.8. Protocol, registration and analytical pre-specification

No a priori study protocol was registered with a public registry. Public protocol registration is not currently mandated by Swiss law, by the ICMJE recommendations or by the relevant reporting guidelines for non-interventional, anonymous professional-opinion surveys; we therefore did not seek registration. The analytical plan — including the pre-specified primary outcome (knowledge-attitude gap, levels 4–5 threshold), the prespecified secondary outcomes, the three sensitivity analyses (knowledge-threshold; non-response Manski bounds; subgroup contrast for current AI users), and the choice of Wilson Score CIs and Cohen’s *h* / *g* effect sizes — was documented internally before data inspection and is fully reproduced in the deposited Python script (Section 2.5; *Data Availability Statement*). No deviations from this internal plan occurred during analysis. Prospective registration of any follow-up surveys within the mediX network is planned.

### 2.9. Ethical considerations

Per Swiss Human Research Act (HFG) Article 2, formal ethics committee approval was not required, as this study collected only professional opinions of physicians without patient data, biological samples or clinical interventions. Participants provided informed consent by proceeding with the voluntary survey after reading the introductory information page, which described the study purpose, the estimated completion time (10–15 minutes), the voluntary and anonymous nature of participation, and the data-handling procedures. Data were analysed in aggregate without individual identification; no IP addresses, names or contact details were retained in the analytical dataset.

### 2.10. Use of generative AI

The author used a commercial large-language-model assistant (Claude, Anthropic) for English-language editing of an earlier manuscript draft and for code review of the analysis script. All scientific content, analytic decisions, interpretation and final wording are the responsibility of the author. No generative AI was used for data fabrication, image generation or quantitative analysis.

## 3. Results

### 3.1. Sample characteristics

Of 620 invited physicians, 160 responded (25.8%); 155 (96.9% of responders) met the pre-specified inclusion criterion of having completed at least two of four core readiness items and were retained for analysis (Supplementary Figure S3). Respondents were distributed across the German-speaking region (n = 137, 88.4%), the Italian-speaking region (Ticino; n = 16, 10.3%) and the French-speaking region (Romandie; n = 2, 1.3%). The age band 40–54 years was modal (n = 71, 45.8%), followed by ≥ 55 years (n = 67, 43.2%) and ≤ 39 years (n = 17, 11.0%); 85 (54.8%) identified as male and 70 (45.2%) as female. Modal years in practice were ≥ 20 (n = 59, 38.1%); the modal practice setting was a group practice with up to three physicians (n = 58, 37.4%). The urbanicity distribution was 63 urban (40.6%), 52 suburban (33.5%), 40 rural (25.8%). Detailed demographics are provided in Supplementary Table S1.

### 3.2. Primary outcome: knowledge–attitude gap

Among 155 respondents, 107/155 (69.0%, 95% CI 61.4–75.8) reported positive attitudes toward AI in primary care (Likert 4–5). In the same 155 respondents, 23/155 (14.8%, 95% CI 10.1–21.3) self-rated their knowledge as high or excellent (categories 4–5). The absolute between-marginal gap was 54.2 percentage points (Figure 1), corresponding to Cohen’s *h* = 1.17 (i.e., > 0.8 large) [14].

**Figure 1.**
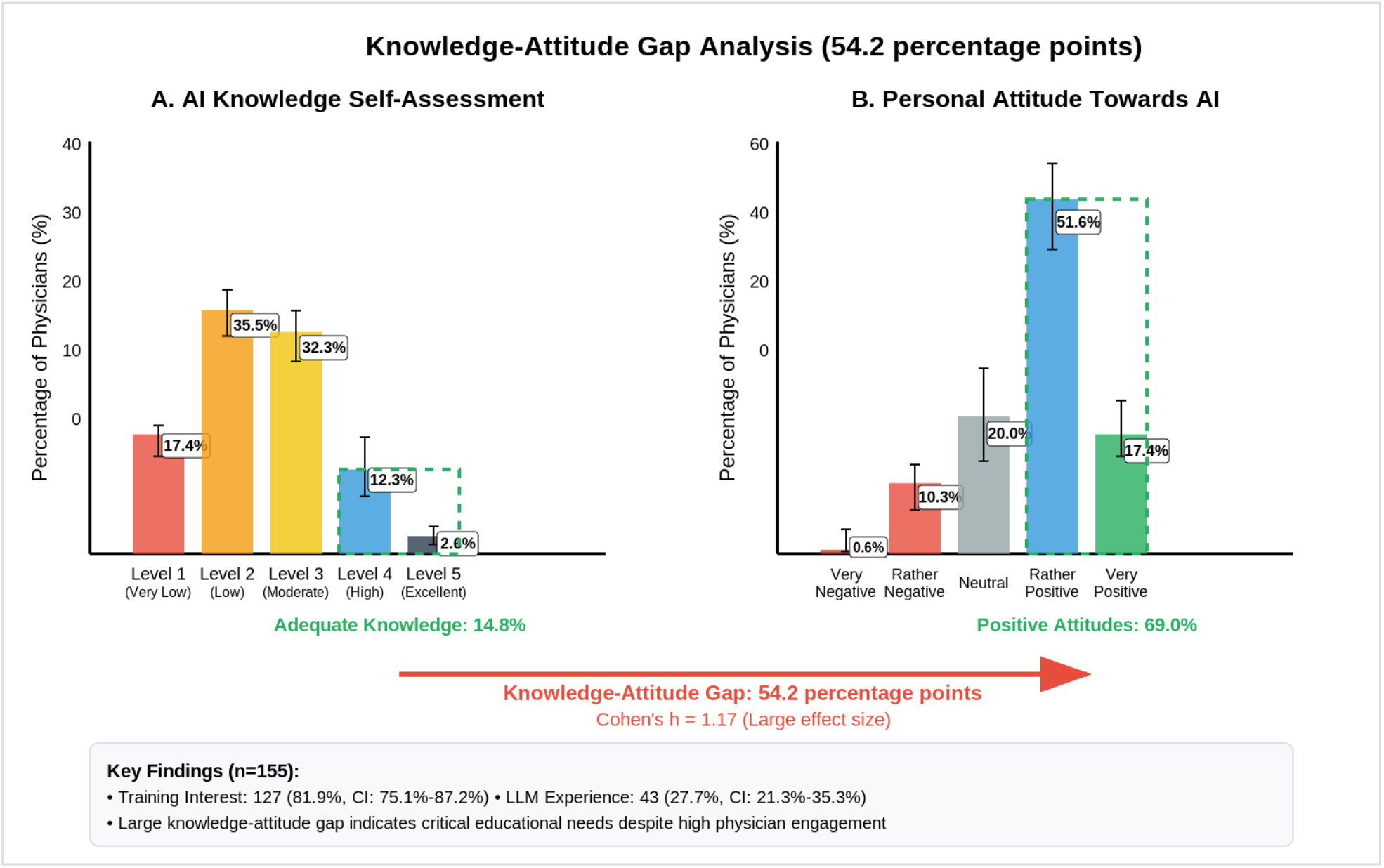
Knowledge–Attitude Gap (n = 155) Distribution of self-rated AI knowledge (Levels 1–5) versus proportion endorsing positive attitudes toward AI (Levels 4–5). Error bars represent 95% Wilson Score confidence intervals. The visualised gap of 54.2 percentage points corresponds to Cohen’s *h* = 1.17 (between-marginal) and Cohen’s *g* = 0.542 (paired McNemar analysis on the same 155 respondents).

Because attitude and knowledge are measured on the same respondents, the comparison is paired and we additionally report an exact McNemar test on the discordant cells of the joint 2 × 2 table. The joint distribution was: positive attitude AND adequate knowledge 20/155 (12.9%); positive attitude only 87/155 (56.1%); adequate knowledge only 3/155 (1.9%); neither 45/155 (29.0%). The exact McNemar test on the 87 vs 3 discordant pairs returned *p* < 0.001; Cohen’s *g* for paired proportions was 0.542 (i.e., > 0.25 large) [14]. 132/155 respondents (85.2%) considered their AI knowledge insufficient (categories 1–3).

### 3.3. Current clinical AI use

43/155 respondents (27.7%, 95% CI 21.3–35.3) reported having already used a large language model (ChatGPT-4, Mistral, Gemini Pro, Anthropic Claude or comparable) for medically-relevant questions and patient care at the time of the survey. Free-text responses identified clinical documentation, drafting of patient information material, decision support and information triage as the principal use cases. Current users held more positive attitudes toward AI than non-users (mean Likert 4.21 vs 3.57; both medians = 4; Mann–Whitney *U* = 1 457, *p* < 0.001; rank-biserial *r* = 0.40 [14]). Current users also self-rated their AI knowledge higher than non-users (mean Likert 3.02 vs 2.26; *U* = 1 421, *p* < 0.001).

### 3.4. Demand for training and framework conditions

127/155 respondents (81.9%, 95% CI 75.1–87.2) endorsed training opportunities (Likert response «Yes, definitely» or «Yes, probably»). Among current AI users this proportion was 39/43 (90.7%, 95% CI 78.4–96.3); among non-users 88/112 (78.6%, 95% CI 70.1–85.2). The most frequently endorsed framework conditions were easy integration into existing workflows 126/155 (81.3%, 95% CI 74.4– 86.6) and sufficient evidence for safety and efficacy 118/155 (76.1%, 95% CI 68.8–82.2). Detailed framework-condition distributions are presented in Supplementary Figure S2.

### 3.5. Implementation priorities

Endorsement rates for the nine prespecified clinical implementation domains are reported in Figure 2 and, with full 95% Wilson Score CIs, in Supplementary Table S2. The three observed support tiers (≥ 70%, 50–69%, < 50%) were not pre-specified analytically; they describe the ex post empirical distribution of endorsement and serve solely as a descriptive structuring device for the eight ranked items (see Section 4.3). Endorsement was 124/155 (80.0%, 95% CI 73.0–85.5) for administrative support and 114/155 (73.5%, 95% CI 66.1–79.9) for image analysis (≥ 70% tier); 100/155 (64.5%) for medication management, 96/155 (61.9%) for diagnostic support and 87/155 (56.1%) for therapy recommendations (50–69% tier); 59/155 (38.1%) for prevention and risk assessment, 43/155 (27.7%) for complex case management, 31/155 (20.0%) for research applications and 26/155 (16.8%) for patient communication (< 50% tier). The between-marginal effect size between the top item (administrative support, 124/155) and the lowest-ranked item (patient communication, 26/155) was Cohen’s *h* = 1.37 [14].

**Figure 2.**
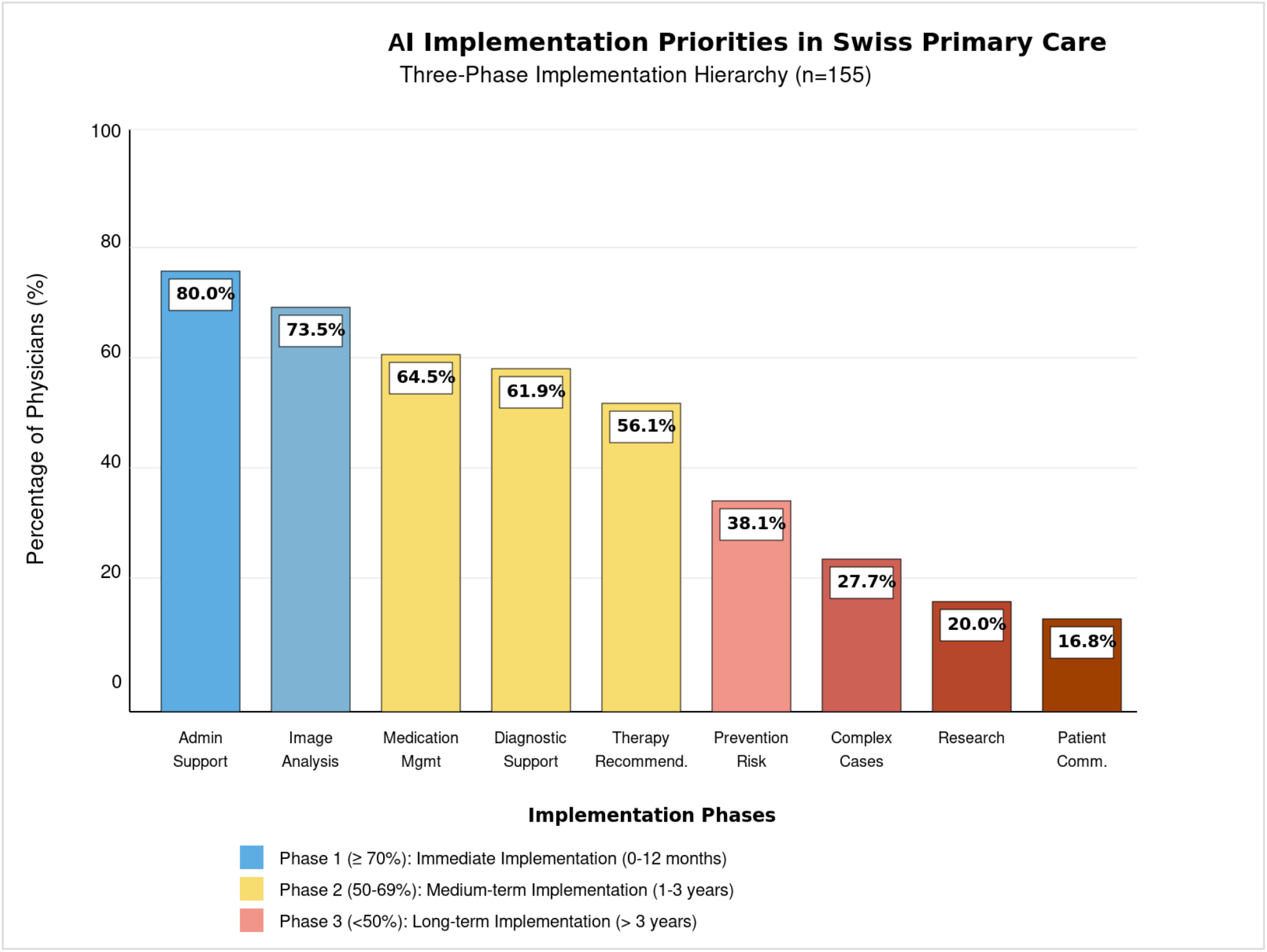
Implementation Priorities — three-tier hierarchy (n = 155) Endorsement of nine prespecified clinical AI implementation domains. Three colour-coded tiers correspond to ≥ 70% endorsement (immediate priority, blue), 50–69% (medium-term, yellow) and < 50% (long-term, red gradient). Bar heights show endorsement percentages; full 95% Wilson Score CIs are reported in Supplementary Table S2 and in Section 3.5.

### 3.6. Perceived benefits and barriers

The leading benefit (administrative relief) was endorsed by 121/155 (78.1%, 95% CI 70.9–83.9) of respondents; the leading barrier (concerns about AI inaccuracy) by 120/155 (77.4%, 95% CI 70.2– 83.3); the between-marginal effect size between these two top-ranked items was Cohen’s *h* = 0.02 [14]. Other top-ranked benefits were efficient workflows 90/155 (58.1%) and continuous monitoring 85/155 (54.8%); other top-ranked barriers were technology dependence 100/155 (64.5%), lack of transparency 87/155 (56.1%) and data protection concerns 77/155 (49.7%). Full benefit and barrier distributions with 95% Wilson Score CIs are presented in Supplementary Figure S1.

### 3.7. Sensitivity analyses

Manski-style worst-case bounds for the population-level training-demand proportion were computed against the sampling frame (n = 620). The lower bound (assuming 0% training demand among the 465 non-responders) was 127/620 (20.5%); the upper bound (assuming 100% training demand among non-responders) was 592/620 (95.5%); the responder-equivalent point estimate (assuming non-responders behave like responders) was 127/155 (81.9%). Under the permissive definition of adequate knowledge (level ≥ 3), 73/155 (47.1%, 95% CI 39.4–55.0) of respondents qualified; the attitude–knowledge between-marginal gap narrowed to 21.9 percentage points (Cohen’s *h* = 0.45 [14]); the corresponding paired McNemar analysis returned *p* < 0.001 with Cohen’s *g* = 0.219, between Cohen 1988 cut-offs for medium (0.15) and large (0.25).

## 4. Discussion

### 4.1. Principal findings

Among 155 primary care physicians from four consenting subnetworks of mediX Switzerland, willingness to adopt clinical AI was high, current clinical use was non-trivial, and self-rated knowledge was low. The 54.2-percentage-point gap between positive attitudes (69.0%) and adequate self-rated knowledge (14.8%) — robust under both a between-marginal Cohen’s *h* analysis and a paired McNemar / Cohen’s *g* analysis, and persistent under a permissive knowledge threshold — describes a population in which conventional resistance-mitigation framings are not the dominant practical concern. The near-equivalent endorsement of the leading benefit (78.1%) and the leading barrier (77.4%) suggests that respondents weigh AI’s potential and its risks symmetrically rather than uncritically.

### 4.2. Comparison with existing literature

The pattern observed here is broadly consistent with international evidence in primary care: a recent systematic review and meta-analysis [6] documents low knowledge alongside positive attitudes across studied populations, and a UK national survey [9] reports that generative AI is in active clinical use among general practitioners despite limited employer encouragement (only 11% encouraged) and minimal formal training (only 5% of users had received AI training). Our findings extend that pattern: in our sample, current AI users were more enthusiastic than non-users yet still self-rated their knowledge as below the high-or-excellent threshold, indicating that early-adoption enthusiasm does not by itself produce competence. Earlier qualitative work from UK primary care [12] anticipated this pattern in terms of physician openness to AI assistance combined with concerns about safety. Reviews of barriers to AI in healthcare more broadly [5,17,18] identify trust, workflow integration, governance and training as recurring themes; our quantitative data corroborate these themes from a Swiss organised-care perspective. The Canadian CFPC AI Working Group report [19] and recent perspectives on AI in graduate medical education [20] both argue, on different evidentiary bases, for systematic readiness assessment before deployment — a recommendation our data support empirically.

### 4.3. The dominant practical risk is unsupervised use, not resistance — and the implementation tiers are descriptive, not prescriptive

The study’s primary contribution is to quantify, within an organised primary care network, a risk profile structurally different from the resistance-based framing that dominates much of the implementation literature: when 27.7% of physicians already use large-language-model tools for medically-relevant work while 85.2% rate their own knowledge as inadequate, the binding constraint on patient safety is not whether physicians will adopt AI but whether their existing adoption is anchored in sufficient understanding of AI failure modes, hallucination, model drift and data-protection requirements. Two methodological caveats accompany this contribution. First, the three-tier classification of implementation priorities (≥ 70% / 50–69% / < 50%) reported in Section 3.5 was derived ex post from the empirical distribution of endorsement and is not an a priori prediction of the «right» order; the cut-offs serve solely as a transparent descriptive structuring device, and any single threshold would yield a similar ordinal ranking. Second, the conclusion that the dominant practical risk is unsupervised use rather than resistance is conditional on the studied population (AI-engaged physicians within the four consenting subnetworks); generalisation to physicians who systematically declined to participate or to non-mediX networks requires additional evidence. The European Union AI Act [7] and the World Health Organization guidance on large multi-modal models [8] both presuppose deployment alongside structured governance; our data suggest that, in primary care, deployment is already running ahead of governance. Educational interventions ranging from short structured continuing-medical-education modules to longer integration into postgraduate curricula [20,21] therefore appear urgent rather than aspirational. From a bio-psycho-social standpoint, the implications extend beyond technical safety: clinicians who use systems they do not feel competent to use are exposed to a particular form of moral and cognitive strain that, if unaddressed, has a plausible link to broader patterns of digital-work-related distress.

### 4.4. Strengths and limitations

The main strengths of this study are: a multilingual sampling frame spanning the three principal Swiss linguistic regions; reporting compliant with CHERRIES and STROBE; pre-specified Wilson Score confidence intervals and effect sizes throughout; transparent non-response and threshold sensitivity analyses; and full availability of the dataset and analysis code under CC-BY-4.0.

Limitations are substantive and the study is therefore framed as exploratory. *First*, the response rate (25.8%) is modest; selection bias toward AI-engaged physicians is *likely but not empirically quantifiable from the available data*, since comparative demographics on non-responders within the consenting subnetworks were not collected. The population-level estimates are best read against the Manski bounds reported in Section 3.7. *Second*, the principal knowledge measure is self-rated and not externally validated; given the well-replicated tendency for self-rated competence in fast-moving technical domains to over-rather than under-estimate true competence, the observed gap is plausibly conservative. *Third*, the four consenting subnetworks (mediX Zurich, Bern, Romandie, Ticino) cover the largest regional units of mediX Switzerland but represent only ≈ 70% of the overall network; smaller subnetworks were not approached and may differ. Generalisation beyond similar organised primary care contexts therefore requires caution. *Fourth*, the cross-sectional design cannot adjudicate causal direction between attitude, use and knowledge; longitudinal designs would be required for that purpose. *Fifth*, the study was conducted by a single investigator without external funding; logistical support and brief consultative input from senior colleagues are acknowledged below. *Sixth*, no a priori study protocol was registered with a public registry (Section 2.8); while public registration is not currently mandated for non-interventional opinion surveys, it would strengthen future iterations of this work.

*Seventh*, the data were collected in August–September 2024 in what is acknowledged to be a rapidly evolving field. Several material developments have occurred between data collection and submission: the European Union AI Act came into force in February 2025 [7]; commercial clinical AI tools have been progressively deployed in Swiss primary care, including embedded large-language-model assistants in electronic health records and clinical-documentation aids; and structured continuing-medical-education modules on AI literacy have begun to proliferate within Swiss postgraduate training. The absolute proportions reported here are therefore best read as a documented baseline rather than as current point estimates — most plausibly, current LLM use is likely higher than 27.7%, and self-rated knowledge somewhat improved relative to the 14.8% level observed in 2024. The structural pattern of the knowledge–attitude gap, the unsupervised-use phenomenon, and the implementation-priority hierarchy are, however, expected to be more durable than the underlying numerical levels, and follow-up surveys within the same network are planned to quantify trajectory and validate the persistence of these patterns under post-AI-Act conditions.

### 4.5. Implications for practice and research

For practice, the data argue for educational and governance scaffolding to be put in place *before* further clinical AI deployment in coordinated primary care, with a sequencing that follows physician preference: administrative and image-analysis applications first; medication and diagnostic support next; complex case management and patient communication last and only after substantive evaluation. For research, three lines follow naturally: a longitudinal re-survey within the same network to quantify trajectory; a randomised evaluation of a structured short-format AI-literacy module embedded in continuing medical education; and a parallel qualitative study to characterise the lived experience of physicians who use AI tools they consider themselves under-prepared to use.

## 5. Conclusions

In a Swiss multilingual primary care network, AI enthusiasm and ongoing clinical AI use coexist with low self-rated competence and minimal training. The dominant practical risk is therefore unsupervised adoption rather than resistance; educational and governance interventions appear required to precede further deployment.

## Abbreviations

AI: artificial intelligence
CHERRIES: Checklist for Reporting Results of Internet E-Surveys
CI: confidence interval
mediX: cooperative network of Swiss primary care practices
STROBE: Strengthening the Reporting of Observational Studies in Epidemiology

## Author Contributions

MV is the sole author and is responsible for all aspects of this work, including conceptualization, methodology, software, formal analysis, investigation, data curation, writing — original draft preparation, writing — review and editing, visualization, project administration and final approval of the submitted version. The author has read and agreed to the published version of the manuscript.

## Institutional Review Board Statement

Per Swiss Human Research Act (Humanforschungsgesetz, HFG) Article 2, formal ethics committee approval was not required as this study collected only professional opinions of physicians without patient data, biological samples or clinical interventions. No application to a Cantonal Ethics Committee was therefore filed.

## Informed Consent Statement

Participants provided informed consent by proceeding with the voluntary online survey after reading the introductory information page describing the study purpose, the estimated completion time (10– 15 minutes), the voluntary and anonymous nature of participation, and the data-handling procedures. Data were analysed in aggregate without individual identification; no IP addresses, names or contact details were retained in the analytical dataset.

## Data Availability Statement

The fully de-identified study dataset, the accompanying data dictionary, the survey instrument in three languages, the CHERRIES and STROBE checklists, and the Python analysis script (v11/v12) are openly available at Zenodo, DOI 10.5281/zenodo.20081687, under a Creative Commons Attribution 4.0 International (CC BY 4.0) licence as of 8 May 2026, indefinitely. The deposited materials cover all variables underlying every reported analysis (155 respondents × 98 retained variables in the public CSV; 104 columns including survey-platform metadata in the original xlsm export). No data-use application is required: any researcher may reuse the materials for any analytic purpose, including derivative or replication work, with attribution. Questions relating to the dataset can be addressed to the corresponding author.

## Acknowledgments

The author wishes to express sincere appreciation to all the primary care physicians from mediX Zurich, mediX Bern, mediX Ticino and mediX Romandie who took the time to participate in this survey. Their willingness to share their professional perspectives and clinical experiences during a period of high practice workload was indispensable to this research and contributes substantively to our collective understanding of artificial intelligence implementation in Swiss primary care.

The author also extends sincere gratitude to the leadership of mediX Switzerland for enabling this study within the network. Their organizational support was essential for the success of the project and represents a meaningful contribution to the development of evidence-based primary healthcare.

Particular thanks are due to **Prof. Corinne Chmiel**, Head of Research at mediX Switzerland, for facilitating the distribution of the survey within the network; and to **Prof. Oliver Senn**, Head of Research and Deputy Director of the Institute of Primary Care at the University of Zurich, for collegial consultative input during the early phase of study design.

The author acknowledges the use of artificial-intelligence tools (Claude, Anthropic) for literature-review assistance, English-language editing and code review. All data analysis, interpretation and scientific conclusions remain under the full oversight and responsibility of the author.

## Supplementary Materials

**Supplementary Figure S1.**
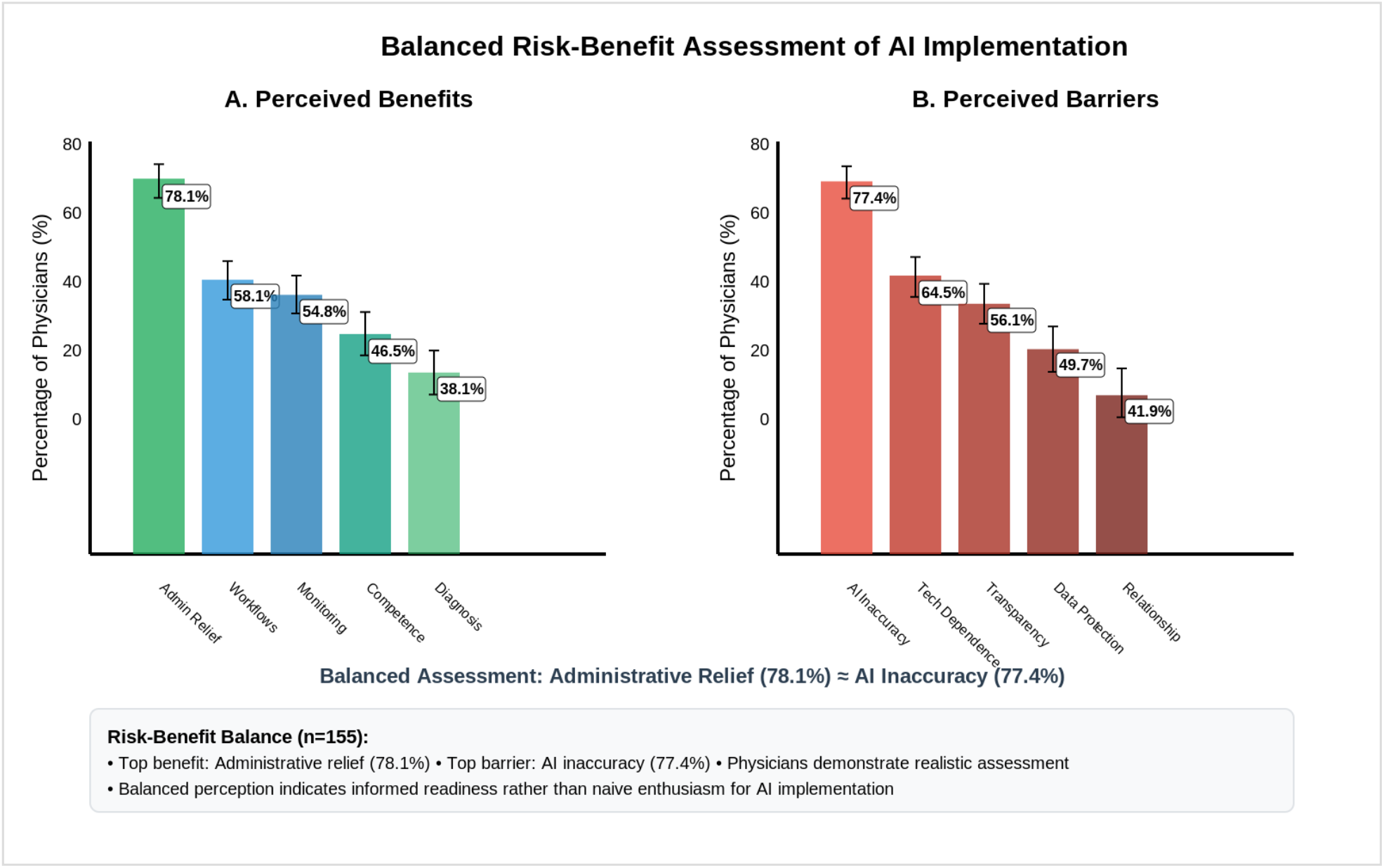
Perceived Benefits and Barriers (n = 155) Top five perceived benefits (left) and top five perceived barriers (right). Error bars represent 95% Wilson Score CIs. The leading benefit (administrative relief, 78.1%) and the leading barrier (AI inaccuracy, 77.4%) are near-equivalent (Cohen’s *h* = 0.02).

**Supplementary Figure S2.**
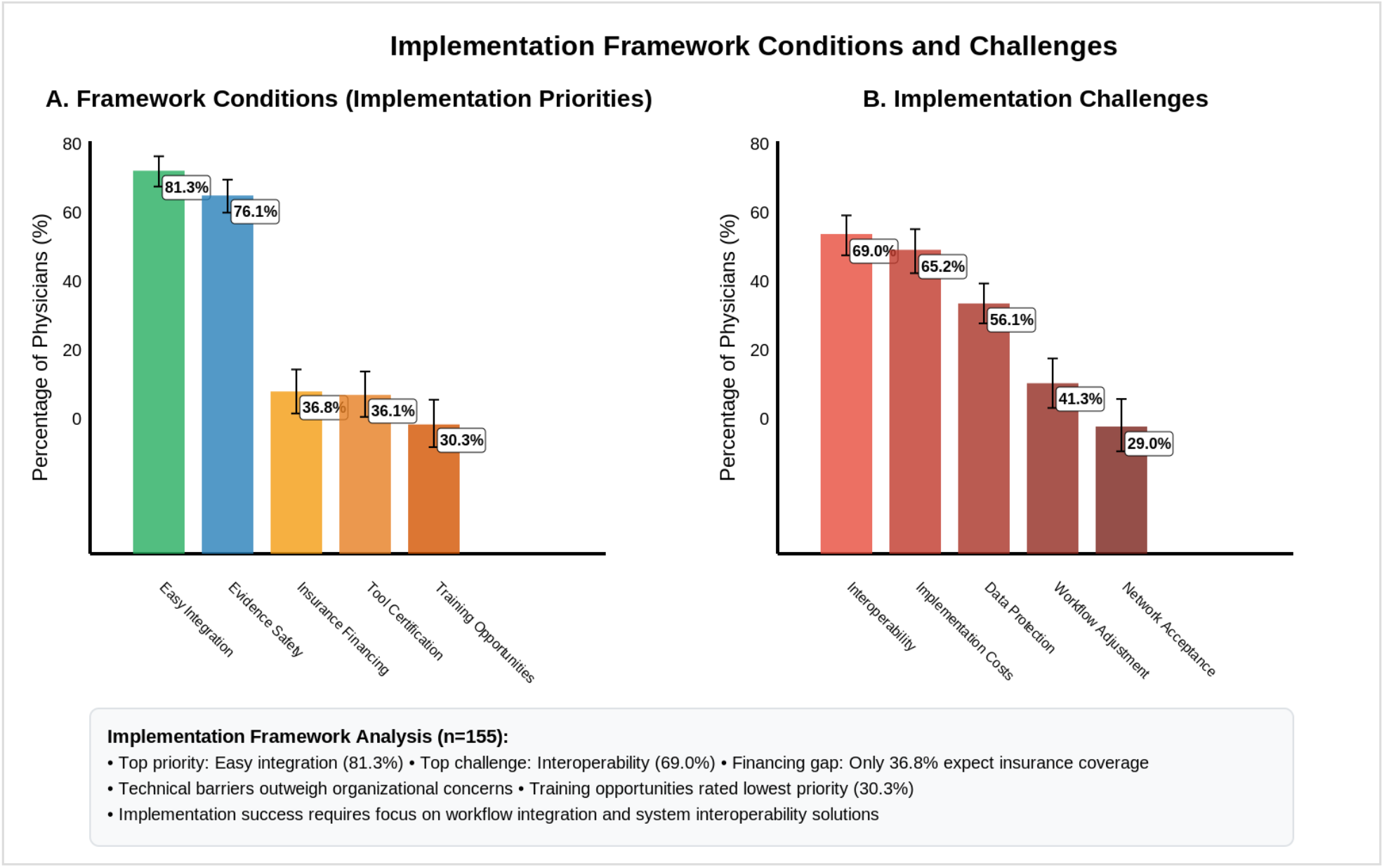
Framework Conditions and Anticipated Implementation Challenges (n = 155) Required framework conditions (left) and anticipated implementation challenges (right). Error bars represent 95% Wilson Score CIs.

**Supplementary Figure S3.**
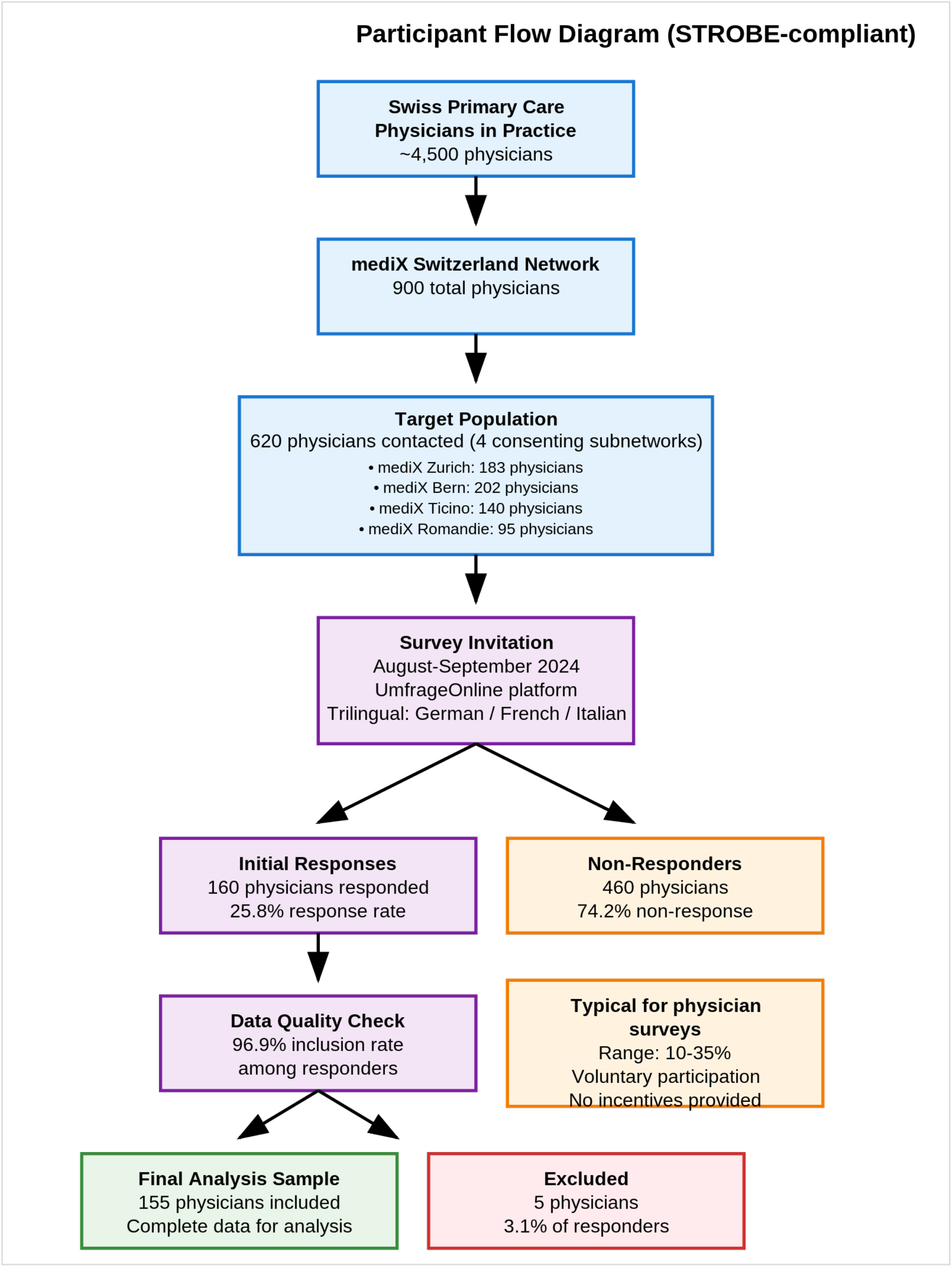
Participant Flow Diagram (STROBE-compliant) Participant flow from sampling frame (n = 620 across four consenting subnetworks: mediX Zurich 183, mediX Bern 202, mediX Ticino 140, mediX Romandie 95) through invitation, response (n = 160; 25.8%), inclusion criterion (≥ 2 of 4 readiness items completed) and final analytical sample (n = 155; 96.9% of responders). Trilingual deployment in German, French and Italian.

## Supplementary Tables

**Supplementary Table S1.** Sample characteristics (n = 155)

| Variable | Category | n / N | % | 95% Wilson CI | Missing |
| --- | --- | --- | --- | --- | --- |
| **Gender** | Male | 85 / 155 | 54.8% | 47.0–62.5 | 0 |
|  | Female | 70 / 155 | 45.2% | 37.5–53.0 | 0 |
| **Language region** | German-speaking | 137 / 155 | 88.4% | 82.4–92.5 | 0 |
|  | Italian-speaking (Ticino) | 16 / 155 | 10.3% | 6.5–16.1 | 0 |
|  | French-speaking (Romandie) | 2 / 155 | 1.3% | 0.4–4.6 | 0 |
| **Age band** | $\leq 39$ years | 17 / 155 | 11.0% | 7.0–16.9 | 0 |
|  | 40–54 years | 71 / 155 | 45.8% | 38.2–53.7 | 0 |
| | $\geq 55$ years | 67 / 155 | 43.2% | 35.7–51.1 | 0 |
| **Years of experience** | < 5 years | 17 / 155 | 11.0% | 7.0–16.9 | 0 |
|  | 5–10 years | 27 / 155 | 17.4% | 12.3–24.2 | 0 |
|  | 10–20 years | 52 / 155 | 33.5% | 26.6–41.3 | 0 |
|  | > 20 years | 59 / 155 | 38.1% | 30.8–45.9 | 0 |
| **Practice setting** | Urban | 63 / 155 | 40.6% | 33.2–48.5 | 0 |
|  | Suburban | 52 / 155 | 33.5% | 26.6–41.3 | 0 |
|  | Rural | 40 / 155 | 25.8% | 19.6–33.2 | 0 |
| **Practice size** | Solo practice | 20 / 155 | 12.9% | 8.5–19.1 | 0 |
|  | Group practice (2–3 physicians) | 58 / 155 | 37.4% | 30.2–45.3 | 0 |
|  | Group practice (4–9 physicians) | 52 / 155 | 33.5% | 26.6–41.3 | 0 |
| | Group practice ($\geq 10$ physicians) | 25 / 155 | 16.1% | 11.2–22.7 | 0 |

*Wilson Score 95% confidence intervals for proportions (Wilson 1927). Listwise missing reported per variable; no imputation performed. $n$ = number of respondents in category; $N$ = analytic sample size. Computed from the deposited dataset (Zenodo, [DOI 10.5281/zenodo.20081687](https://doi.org/10.5281/zenodo.20081687)) using the deposited Python analysis script (v11/v12).*

**Supplementary Table S2.** Full implementation-priority ranking with 95% Wilson Score CIs (n = 155)

| Rank | Implementation domain | n / N | % endorsement | 95% Wilson CI | Support tier |
| --- | --- | --- | --- | --- | --- |
| 1 | Administrative support | 124 / 155 | 80.0% | 73.0–85.5 | ≥ 70% (immediate) |
| 2 | Image analysis | 114 / 155 | 73.5% | 66.1–79.9 | ≥ 70% (immediate) |
| 3 | Medication management | 100 / 155 | 64.5% | 56.7–71.6 | 50–69% (medium-term) |
| 4 | Diagnostic support | 96 / 155 | 61.9% | 54.1–69.2 | 50–69% (medium-term) |
| 5 | Therapy recommendations | 87 / 155 | 56.1% | 48.3–63.7 | 50–69% (medium-term) |
| 6 | Prevention and risk assessment | 59 / 155 | 38.1% | 30.8–45.9 | < 50% (long-term) |
| 7 | Complex case management | 43 / 155 | 27.7% | 21.3–35.3 | < 50% (long-term) |
| 8 | Research applications | 31 / 155 | 20.0% | 14.5–27.0 | < 50% (long-term) |
| 9 | Patient communication | 26 / 155 | 16.8% | 11.7–23.4 | < 50% (long-term) |

*Endorsement counts respondents who selected the item when asked to choose at most five priorities from nine prespecified clinical AI implementation domains. Wilson Score 95% CIs (Wilson 1927). The three-tier classification (≥ 70% / 50–69% / < 50%) was derived ex post from the empirical endorsement distribution and serves as a descriptive structuring device, not as an a priori prediction (see Section 4.3). The between-marginal effect size between the top item (administrative support, 124/155) and the lowest-ranked item (patient communication, 26/155) is Cohen's $h = 1.37$ [14].*

## References

1. American Medical Association. AI Usage Among Doctors Doubles as Confidence in Technology Grows; AMA: Chicago, IL, USA, 2024. Available online: https://www.ama-assn.org/press-center/ama-press-releases/ama-ai-usage-among-doctors-doubles-confidence-technology-grows (accessed on 7 May 2026).

2. Topol, E.J. High-performance medicine: The convergence of human and artificial intelligence. Nat. Med. 2019, 25, 44–56. 10.1038/s41591-018-0300-7.

3. Moor, M.; Banerjee, O.; Abad, Z.S.H.; Krumholz, H.M.; Leskovec, J.; Topol, E.J.; Rajpurkar, P. Foundation models for generalist medical artificial intelligence. Nature 2023, 616, 259–265. 10.1038/s41586-023-05881-4.

4. Nori, H.; King, N.; McKinney, S.M.; Carignan, D.; Horvitz, E. Capabilities of GPT-4 on Medical Challenge Problems. arXiv 2023, arXiv:2303.13375. 10.48550/arXiv.2303.13375.

5. Hassan, M.; Kushniruk, A.; Borycki, E. Barriers to and Facilitators of Artificial Intelligence Adoption in Health Care: Scoping Review. JMIR Hum. Factors 2024, 11, e48633. 10.2196/48633.

6. Abdulazeem, H.M.; Meckawy, R.; Schwarz, S.; Novillo-Ortiz, D.; Klug, S.J. Knowledge, attitude, and practice of primary care physicians toward clinical AI-assisted digital health technologies: Systematic review and meta-analysis. Int. J. Med. Inform. 2025, 201, 105920. 10.1016/j.ijmedinf.2025.105920.

7. Busch, F.; Kather, J.N.; Johner, C.; Moser, M.; Truhn, D.; Adams, L.C.; Bressem, K.K. Navigating the European Union Artificial Intelligence Act for healthcare. NPJ Digit. Med. 2024, 7, 210. 10.1038/s41746-024-01213-6.

8. World Health Organization. Ethics and Governance of Artificial Intelligence for Health: Guidance on Large Multi-Modal Models; WHO: Geneva, Switzerland, 2024; ISBN 978-92-4-008475-9.

9. Kharko, A.; Garcia Sanchez, C.; Hagström, J.; Gaab, J.; Locher, C.; McMillan, B.; Sundemo, D.; Blease, C. General practitioners’ opinions of generative artificial intelligence in the UK: An online survey. Digit. Health 2025, 11, 20552076251360863. 10.1177/20552076251360863.

10. Amon, C.; King, J.; Colclasure, J.; Hodge, K.; DuBard, C.A. Leveraging Accountable Care Organization infrastructure for rapid pandemic response in independent primary care practices. Healthc. (Amst.) 2022, 10, 100640. 10.1016/j.hjdsi.2022.100640.

11. Venkatesh, V.; Morris, M.G.; Davis, G.B.; Davis, F.D. User acceptance of information technology: Toward a unified view. MIS Q. 2003, 27, 425–478. 10.2307/30036540.

12. Blease, C.; Kaptchuk, T.J.; Bernstein, M.H.; Mandl, K.D.; Halamka, J.D.; DesRoches, C.M. Artificial Intelligence and the Future of Primary Care: Exploratory Qualitative Study of UK General Practitioners’ Views. J. Med. Internet Res. 2019, 21, e12802. 10.2196/12802.

13. Wilson, E.B. Probable inference, the law of succession, and statistical inference. J. Am. Stat. Assoc. 1927, 22, 209–212. 10.2307/2276774.

14. Cohen, J. Statistical Power Analysis for the Behavioral Sciences, 2nd ed.; Lawrence Erlbaum Associates: Hillsdale, NJ, USA, 1988; ISBN 978-0-8058-0283-2.

15. Eysenbach, G. Improving the quality of web surveys: The Checklist for Reporting Results of Internet E-Surveys (CHERRIES). J. Med. Internet Res. 2004, 6, e34. 10.2196/jmir.6.3.e34.

16. von Elm, E.; Altman, D.G.; Egger, M.; Pocock, S.J.; Gøtzsche, P.C.; Vandenbroucke, J.P.; STROBE Initiative. The Strengthening the Reporting of Observational Studies in Epidemiology (STROBE) statement: Guidelines for reporting observational studies. Lancet 2007, 370, 1453–1457. 10.1016/S0140-6736(07)61602-X.

17. Nair, M.; Svedberg, P.; Larsson, I.; Nygren, J.M. A comprehensive overview of barriers and strategies for AI implementation in healthcare: Mixed-method design. PLoS ONE 2024, 19, e0305949. 10.1371/journal.pone.0305949.

18. Kelly, C.J.; Karthikesalingam, A.; Suleyman, M.; Corrado, G.; King, D. Key challenges for delivering clinical impact with artificial intelligence. BMC Med. 2019, 17, 195. 10.1186/s12916-019-1426-2.

19. Kueper, J.K.; Emu, M.; Banbury, M.; Bjerre, L.M.; Choudhury, S.; Green, M.; Pimlott, N.; Slade, S.; Tsuei, S.H.; Sisler, J. Artificial intelligence for family medicine research in Canada: Current state and future directions: Report of the CFPC AI Working Group. Can. Fam. Physician 2024, 70, 161–168.

20. College of Family Physicians of Canada. Artificial Intelligence in Graduate Medical Education. Ann. Fam. Med. 2025, 23, 480–481. 10.1370/afm.250512.

21. Hoffman, M. Competency-based and less time-bound: A new approach to the macro-structure of a medical school curriculum. Med. Educ. Online 2024, 29, 2343205. 10.1080/10872981.2024.2343205.

