## Supplementary material for "Knowledge–Attitude Gap and Unsupervised Use of Artificial Intelligence Tools Among Swiss Primary Care Physicians: A Multicentric Cross-Sectional Survey": Supplementary_File_S1_CHERRIES_Checklist.pdf

### CHERRIES Checklist v4 — *Healthcare* (MDPI), Special Issue submission

**Checklist for Reporting Results of Internet E-Surveys (CHERRIES, Eysenbach 2004)**

**Study:** Knowledge–Attitude Gap and Unsupervised Use of Artificial Intelligence Tools Among Swiss Primary Care Physicians: A Multicentric Cross-Sectional Survey **Author:** Marco Vecellio, MD — ORCID 0009-0000-1772-5620 **Date:** 8 May 2026 **Manuscript version:** v11 / Skript-Version v11(v12) **Submission target:** *Healthcare* (MDPI), Special Issue «Artificial Intelligence in Healthcare: Opportunities and Challenges» — Invitation 119530958/OVBEJco5 **Survey period:** 1 August – 30 September 2024

#### 1. Design

**Item 1: Describe survey design**

> Cross-sectional, descriptive, web-based survey of primary care physicians within mediX Switzerland, deployed via the professional online platform UmfrageOnline. The design is best characterised as a **closed-network, census-style invitation with self-selected respondents**: the entire eligible frame of 620 physicians within the four consenting subnetworks (mediX Zurich, Bern, Romandie, Ticino — jointly representing the largest regional units across all three principal Swiss linguistic regions) received the same survey invitation; participation was voluntary, anonymous and uncompensated. The instrument was deployed in three languages (German, French, Italian) to mirror Swiss linguistic diversity.

#### 2. IRB Approval and Informed Consent

**Item 2: IRB approval**

> Per Swiss Human Research Act (Humanforschungsgesetz, HFG) Article 2, formal ethics committee approval was not required, as this study collected only professional opinions of physicians without patient data, biological samples or clinical interventions. No application to a Cantonal Ethics Committee was therefore filed. Data protection measures included full anonymisation in the analytical dataset, secure data storage on the survey-platform infrastructure, encrypted transmission, access controls and retention limitations in compliance with Swiss data protection regulations.

**Item 3: Informed consent**

> Participants were informed about the study purpose, the estimated completion time (10–15 minutes), the voluntary and anonymous nature of participation, and the data-handling procedures via the survey landing page. Informed consent was obtained electronically by proceeding with the survey after reading the information page. Data were analysed in aggregate without individual identification.

#### 3. Data Collection Process

**Item 4: Data collection mode**

> Web-based survey administered via UmfrageOnline ( `https://www.umfrageonline.com` ). The survey was accessible via a direct invitation link distributed through the four consenting subnetworks' internal communication infrastructure (HIN secure professional email).

**Item 5: Duplicate entries**

> Duplicate submissions were prevented by the platform's session management and IP-address checking; the analytical dataset retains no IP information.

**Item 6: Development and testing**

> The survey instrument was developed by the author from existing literature on physician AI attitudes and adapted to the Swiss context. The questionnaire was pilot-tested with **five primary care physicians** (not subsequently invited to the main study) and refined accordingly; three items with ambiguous wording were revised after pilot. Content review was conducted with consultative input from Prof. Oliver Senn (University of Zurich, Institute of Primary Care) and Prof. Corinne Chmiel (mediX Zurich).

#### 4. Recruitment and Sample Description

**Item 7: Open survey vs closed survey**

> Closed survey. Access was restricted to physicians within the four consenting mediX subnetworks via direct invitation link.

**Item 8: Contact mode**

> Participants were contacted through the secure professional email infrastructure of the four consenting subnetworks (HIN email). The survey invitation was distributed via the subnetworks' established physician communication channels with two reminder emails at two-week intervals.

**Item 9: Advertising the survey**

> The survey was announced exclusively within the four consenting mediX subnetworks. No external advertising or public promotion was conducted.

#### 5. Survey Administration

**Item 10: Web/E-mail**

> Web-based survey administered through UmfrageOnline; invitation by HIN email.

**Item 11: Context**

> The survey was presented within the context of AI implementation research in primary care. Participants accessed the survey through the UmfrageOnline portal, clearly identified as a research initiative within their professional mediX subnetwork.

**Item 12: Mandatory/voluntary**

> Voluntary participation. All survey items were optional, and participants could skip questions or discontinue participation at any time without consequences.

**Item 13: Incentives**

> No financial or material incentives were provided. Participation was based on professional interest in AI implementation research.

**Item 14: Time/Date**

> The survey was open from 1 August 2024 to 30 September 2024. Reminder communications were sent twice during the survey period (after 14 and 28 days).

**Item 15: Randomization of items or questionnaires**

> No randomisation of survey items was implemented; all participants received the same items in the same order to ensure consistency and comparability.

**Item 16: Adaptive questioning**

> No adaptive or conditional questioning was used; all participants were presented with the same set of items regardless of their previous responses (apart from automatic linguistic routing among the German, French and Italian instruments).

**Item 17: Number of items**

> The instrument comprised **35 substantive items** distributed across nine sections: demographic and professional characteristics; AI knowledge self-assessment; AI attitudes; current LLM use; perceived benefits; perceived barriers; implementation priorities; framework conditions; demand for training. The raw export from the survey platform contains 104 columns including platform metadata.

**Item 18: Number of screens (pages)**

> The survey was presented across multiple screens to improve user experience; participants could navigate between pages and review their responses before final submission.

**Item 19: Completeness check**

> The platform included a review function allowing participants to check their responses before final submission; a final "Submit" button was required to complete the survey.

**Item 20: Review step**

> Yes; participants could return to previous pages to modify responses before final submission.

#### 6. Response Rates

**Item 21: Unique site visitors**

> Unique site visitors were tracked by the survey platform. Of 620 invited physicians, 160 unique visitors accessed and submitted to the survey.

**Item 22: View rate**

> View rate is not reported as a separate metric for this closed survey, since the invitation link was sent only to the 620 eligible network members; we report participation and completion rates instead.

**Item 23: Participation rate**

> Participation rate: **160/620 (25.8%)**.

**Item 24: Completion rate**

> Completion rate among participants: **155/160 (96.9%)** with respect to the inclusion criterion of completing ≥ 2 of 4 core readiness items.

#### 7. Preventing Multiple Entries

**Item 25: Cookies used**

> Standard session cookies were used by the UmfrageOnline platform for navigation and to prevent in-session double submission. Cookies were not used for persistent identification; no identifying cookie data are retained in the analytical dataset.

**Item 26: IP checking**

> IP-address checking was implemented at the platform level to prevent duplicate submissions. The analytical dataset retains no IP information (the IP-address column is uniformly anonymised to "-" in the source export and stripped entirely from the public CSV).

**Item 27: Log file analysis**

> Platform access logs supported the calculation of participation and completion rates. Logs were not retained beyond the analytical period.

**Item 28: Registration**

> Registration with a public study registry was not undertaken. Public registration is not currently mandated for non-interventional anonymous opinion surveys of healthcare professionals; the

> analytical plan was documented internally before data inspection and is fully reproduced in the deposited Python script (Zenodo, DOI 10.5281/zenodo.20081687).

#### 8. Analysis

**Item 29: Handling of incomplete questionnaires**

> Incomplete questionnaires were handled by listwise deletion: of 160 initial responses, 5 were excluded for failing the inclusion criterion (≥ 2 of 4 core readiness items completed), yielding a final analytical sample of 155 (96.9% inclusion rate). Per-item denominators are reported alongside each estimate; missing data on individual non-readiness items were minimal (< 5%).

**Item 30: Atypical timestamps**

> No atypical timestamps were identified. All submissions occurred within the designated survey period.

**Item 31: Statistical correction**

> No formal multiple-testing corrections were applied, given the exploratory descriptive nature of the study. Wilson Score 95% confidence intervals were used throughout for proportions; effect sizes were quantified using Cohen's *h* for between-marginal proportion contrasts and Cohen's *g* for paired-proportion contrasts (Cohen 1988); the principal paired comparison (knowledge–attitude gap) was additionally tested with an exact McNemar test. Manski-style worst-case bounds against the sampling frame were computed for the population-level training-demand estimate. Software: Python 3.11 with pandas 2.1, scipy 1.11, numpy 1.26 and openpyxl 3.1 (analysis script v11/v12 deposited at Zenodo under CC BY 4.0).

---

#### Summary

This CHERRIES checklist demonstrates conformity of the survey methodology with the Eysenbach 2004 standard, with full transparency on response rates, technical safeguards, ethical framing under HFG Art. 2, and analytical reproducibility through deposited dataset and code.

**Key quality indicators**

- Closed-network, census-style invitation of all 620 eligible physicians in four consenting subnetworks.
- Voluntary, anonymous, uncompensated participation.
- IP-based duplicate prevention; analytical dataset fully de-identified.
- Participation rate 25.8% (160/620); completion rate 96.9% (155/160).
- Pilot-tested instrument (5 physicians); three languages (German, French, Italian).
- Deposited dataset, codebook, instrument and analysis script under CC BY 4.0.

**Key updated metrics (manuscript v11)**

- Knowledge–Attitude Gap: 54.2 percentage points (Cohen's $h$ = 1.17 between-marginal; Cohen's $g$ = 0.542 paired; McNemar exact $p < 0.001$).
- Current LLM use: 27.7% (95% CI 21.3–35.3).
- Training demand: 81.9% (95% CI 75.1–87.2); Manski bounds 20.5% – 95.5%.
- Top implementation priority — Administrative support: 80.0% (95% CI 73.0–85.5).

**Prepared for:** *Healthcare* (MDPI) Special Issue submission, May 2026. **Compliance:** Full CHERRIES checklist completion. **Version:** v4 — 8 May 2026.
