## Supplementary material for "Knowledge–Attitude Gap and Unsupervised Use of Artificial Intelligence Tools Among Swiss Primary Care Physicians: A Multicentric Cross-Sectional Survey": Supplementary_File_S2_STROBE_Checklist.pdf

### STROBE Checklist v4 — *Healthcare* (MDPI), Special Issue submission

**STROBE Statement — Checklist of items that should be included in reports of observational studies (cross-sectional)**

**Study Title:** Knowledge–Attitude Gap and Unsupervised Use of Artificial Intelligence Tools Among Swiss Primary Care Physicians: A Multicentric Cross-Sectional Survey **Author:** Marco Vecellio, MD — ORCID 0009-0000-1772-5620 **Journal:** *Healthcare* (MDPI), Special Issue «Artificial Intelligence in Healthcare: Opportunities and Challenges» **Submission Reference:** Invitation 119530958/OVBEJco5 **Date:** 8 May 2026 **Version:** v4 — Healthcare submission

| Item | Recommendation | Section / Page | Status | Comments |
|---|---|---|---|---|
| **Title and abstract** | | | | |
| 1 (a) | Indicate the study's design with a commonly used term in the title or the abstract | Title; Abstract §1 | ✓ | «Cross-Sectional Survey» in title; «closed-network, census-style online survey» in Abstract §Methods. |
| 1 (b) | Provide in the abstract an informative and balanced summary of what was done and what was found | Abstract | ✓ | Structured abstract: Background / Methods / Results / Conclusions; n/N format throughout. |
| **Introduction** | | | | |
| 2 | Explain the scientific background and rationale for the investigation being reported | §1 | ✓ | AI-implementation acceleration, paradigm-shift framing, organised-care vantage point. |
| 3 | State specific objectives, including any prespecified hypotheses | end of §1 | ✓ | Three explicit aims: (i) quantify gap; (ii) characterise current AI use; (iii) elicit |

| Item | Recommendation | Section / Page | Status | Comments |
|---|---|---|---|---|
| | | | | implementation preferences. |
| **Methods** | | | | |
| 4 | Present key elements of study design early in the paper | §2.1 | ✓ | Cross-sectional, anonymous, web-based; closed-network census-style invitation. |
| 5 | Describe the setting, locations, and relevant dates, including periods of recruitment and exposure | §2.1 | ✓ | mediX Switzerland; four consenting subnetworks; 1 Aug – 30 Sep 2024. |
| 6 (c) Cross-sectional study | Give the eligibility criteria, and the sources and methods of selection of participants | §2.2 | ✓ | All 620 physicians on the four subnetworks' combined HIN distribution lists invited; eligibility = practising mediX primary care physician. **Not convenience sampling**; explicitly classified as closed-network census-style invitation with self-selected respondents. |
| 7 | Clearly define all outcomes, exposures, predictors, potential confounders, and effect modifiers | §2.3, §2.4 | ✓ | Primary outcome (knowledge–attitude gap, level ≥ 4 strict); secondary outcomes (current AI use, training demand, ranked priorities, benefits, barriers). |
| 8 | For each variable of interest, give sources of data and details of methods of assessment (measurement) | §2.3 | ✓ | 5-point Likert scales; multiple-choice domains; pilot-tested with 5 physicians; three-language deployment. |
| 9 | | §2.6 | ✓ | |

| Item | Recommendation | Section / Page | Status | Comments |
|---|---|---|---|---|
| | Describe any efforts to address potential sources of bias | | | Manski-style non-response bounds; threshold sensitivity (level ≥ 4 vs ≥ 3); subgroup contrast (current users vs non-users). |
| 10 | Explain how the study size was arrived at | §2.2 | ✓ | Sampling frame n = 620 (four consenting subnetworks ≈ 70% of full mediX network ≥ 900). |
| 11 | Explain how quantitative variables were handled in the analyses. If applicable, describe which groupings were chosen and why | §2.4, §2.5 | ✓ | Knowledge level ≥ 4 (strict, primary) and ≥ 3 (permissive, sensitivity) explicitly defined; rationale stated. |
| 12 (a) | Describe all statistical methods, including those used to control for confounding | §2.5 | ✓ | Wilson Score 95% CI [13]; Cohen's *h* and *g* (Cohen 1988) [14]; Mann–Whitney *U* with rank-biserial *r*; Spearman ρ; exact McNemar for paired primary outcome. |
| 12 (b) | Describe any methods used to examine subgroups and interactions | §2.6, §3.3 | ✓ | Mann–Whitney *U* contrast for current AI users vs non-users on attitude and knowledge. |
| 12 (c) | Explain how missing data were addressed | §2.5 | ✓ | Listwise deletion at participant level; per-item denominators reported; no imputation. |
| 12 (f) Cross-sectional | Describe analytical methods taking account of sampling strategy | §2.6 | ✓ | Manski-style non-response bracketing computed against |

| Item | Recommendation | Section / Page | Status | Comments |
|---|---|---|---|---|
| | | | | the sampling frame (n = 620). |
| 13 | Describe any sensitivity analyses | §2.6 | ✓ | Three sensitivity analyses pre-specified: knowledge threshold; non-response bounds; subgroup contrast. |
| **Methods (additional, study-specific)** | | | | |
| Open Science — Protocol & registration | §2.8 | ✓ | No a priori registered protocol; analytical plan internally documented before data inspection and reproduced in deposited Python script. | |
| Open Science — Code & software | §2.5 | ✓ | Python 3.11 with pandas 2.1 (BSD), scipy 1.11 (BSD), numpy 1.26 (BSD), openpyxl 3.1 (MIT), matplotlib 3.8 (PSF); script `analysis_swissAIsurvey_v11.py` (internally v12) at Zenodo, DOI 10.5281/zenodo.20081687, CC BY 4.0. | |
| Ethical considerations | §2.9 | ✓ | Per HFG Art. 2 outside scope of Swiss HRA; no Cantonal Ethics Committee application required and none filed. | |
| **Results** | | | | |
| 14 (a) | Report numbers of individuals at each stage of study | §3.1; Supp. Fig. S3 | ✓ | 620 invited → 160 responded (25.8%) → 155 included (96.9% of responders). |
| 14 (b) | Reasons for non-participation at each stage | §3.1 | ✓ | 5 responders excluded for non-completion of ≥ 2 of 4 core readiness items; non-responder reasons not collected (acknowledged in §4.4 *Limitations*). |
| 14 (c) | Consider use of a flow diagram | Supp. Fig. S3 | ✓ | Participant flow diagram (STROBE-compliant). |
| 15 (a) | | | ✓ | |

| Item | Recommendation | Section / Page | Status | Comments |
|---|---|---|---|---|
| | Give characteristics of study participants | §3.1; Table 1; Supp. Table S1 | | Demographics: 54.8% male; modal age 40–54 (45.8%); modal experience > 20 years (38.1%); 88.4% German-speaking. |
| 15 (b) | Indicate number of participants with missing data for each variable | Table 1 footnote; Supp. Table S1 | ✓ | Per-item denominators reported; missing-data column included in Supp. Table S1. |
| 16 (a) | Give unadjusted estimates and their precision (95% CIs) | §3.2–§3.7; Figures 1–2 | ✓ | All proportions with Wilson Score 95% CIs; effect sizes Cohen's *h* and *g* with Cohen 1988 cut-offs. |
| 16 (b) | Report category boundaries when continuous variables were categorized | §3.2 | ✓ | Knowledge: levels 1–3 insufficient (85.2%), 4–5 adequate (14.8%); attitudes: 1–2 negative (10.9%), 3 neutral (20.0%), 4–5 positive (69.0%). |
| **Discussion** | | | | |
| 17 | Summarise key results with reference to study objectives | §4.1 | ✓ | 54.2-pp gap; 27.7% LLM use; 81.9% training demand; three-tier implementation hierarchy. |
| 18 | Discuss limitations of the study, taking into account sources of potential bias or imprecision | §4.4 | ✓ | Six limitations explicitly named: response rate / self-selection; self-rated knowledge; partial network coverage; cross-sectional design; single-investigator constraint; absence of registered protocol. |

| Item | Recommendation | Section / Page | Status | Comments |
| --- | --- | --- | --- | --- |
| 19 | Give a cautious overall interpretation consistent with results and balancing benefits and harms | §4.1, §4.5 | ✓ | Conditional on AI-engaged subset; balanced benefit–barrier appraisal noted. |
| 20 | Discuss the generalisability (external validity) of the study results | §4.4, §4.5 | ✓ | Generalisation beyond similar organised primary care contexts requires caution; population profile parallels ACO / PCN structures internationally. |
| **Other information** | | | | |
| 21 | Give the source of funding and the role of the funders | *Funding* statement | ✓ | No external funding declared. |
| 22 | (Cohort/Case-control) — n/a | — | n/a | Not applicable for cross-sectional study. |

#### Summary

This STROBE checklist v4 replaces the v3 / BJGP-Open-prepared version and reflects manuscript v11 prepared for *Healthcare* (MDPI). All 22 STROBE items applicable to cross-sectional studies are addressed. Page-number references will be updated at proof stage; the section-number map above is stable.

**Prepared for:** *Healthcare* (MDPI) Special Issue submission, May 2026. **Version:** v4 — 8 May 2026.
